# Exercise intervention for the management of chemotherapy-induced peripheral neuropathy: A systematic review and network meta-analysis

**DOI:** 10.1101/2023.11.13.23298326

**Authors:** Natsuki Nakagawa, Sena Yamamoto, Akiko Hanai, Ayano Oiwa, Harue Arao

## Abstract

Although exercise is recommended for cancer survivors with chemotherapy-induced peripheral neuropathy (CIPN), the effective types of exercise for preventing and treating CIPN remain unclear. This systematic review and network meta-analysis (NMA) aimed to evaluate the comparative effects of exercise on CIPN. We included relevant randomized controlled trials (RCTs) identified in a 2019 systematic review that evaluated the effects of exercise on CIPN and conducted an additional search for RCTs published until 2023. We evaluated the risk of bias for each RCT; the comparative effectiveness of exercise on patient-reported quality of life (QOL) through an NMA; and the effectiveness of exercise on QOL scores, patient-reported CIPN symptoms, and pain through additional meta-analyses.Twelve studies (exercise, n=540; control, n=527) comparing 8 exercise interventions were included in the analysis. All studies were determined to have a high risk of bias. The meta-analyses showed significantly improved QOL (standard mean differences [SMD] 0.45; 95% confidence interval [CI] = 0.12 to 0.78) and CIPN symptoms (SMD 0.46; 95% CI = 0.11 to 0.82). No severe adverse events were reported. Pain tended to improve with exercise (SMD 0.84; 95% CI = −0.11 to 1.80). An NMA suggested that the interventions of combination of balance and strength training showed the significant improvement in QOL scores compared to the control. Exercise interventions are safe and effective in improving QOL. For exercise categories, the combination of balance and strength training would be the promising program. Patients with CIPN will benefit from frequent exercise focusing on the declined symptoms.

## 2 Introduction

Approximately half of adult cancer patients receiving neurotoxic chemotherapy experience chemotherapy-induced peripheral neuropathy (CIPN) (1), which frequently continues after chemotherapy cessation, sometimes for years (1–3). CIPN can result in dose reduction or discontinuation of cancer chemotherapy (4, 5). The rebellious and chronic nature of CIPN can lead to functional and psychiatric impairment, including slower gait and a higher risk of falls compared to cancer patients without CIPN (6–8). Therefore, CIPN can seriously adversely affect quality of life (QOL) (2). It also causes an economic burden due to the long-term prescription of ineffective analgesics for chronic CIPN (9). However, recent clinical guidelines for CIPN indicate that it is difficult to establish effective management of CIPN, except that some patients may benefit from duloxetine as only a partial symptomatic treatment (10, 11).

Previous studies have reported that physical activity and exercise improve cardiovascular fitness, muscle strength, health-related QOL, depression, cachexia, and cancer-related fatigue (12–15). Indeed, the guidelines of the American Society of Clinical Oncology (ASCO) for patients receiving active cancer treatment recommend regular exercise as supportive or palliative care (16).

Although exercise therapy for CIPN mitigation has not been established as part of any established guideline, several systematic reviews imply the potential efficacy of exercise for the prevention and treatment of symptoms related to CIPN (10, 11, 13, 17-19). In these studies, the dose, frequency, intensity, and duration of exercise programs varied (i.e., ranging from inactivity avoidance instructions to resistance training), and the outcome (e.g., specific sensorimotor functions, activities of daily living, QOL) also varied across the studies. According to the ASCO CIPN guidelines, exercise therapy is not currently recommended as a preventative or treatment approach for CIPN because of the low quality of the available evidence based on a systematic review of the literature published between January 2013 and August 2019 (10). The guidelines state that further robust research is necessary to determine the efficacy and potential risks of exercise therapy (10). In a systematic review conducted by Lin et al. (17), the overall mean effect size was estimated using standardized mean differences and a fixed-effect model, and significant improvement in the exercise cohort compared to the control group was reported (mean difference: 0.5319; 95% confidence interval: 0.2295 to 0.8344; Z = 3.45) . However, the study suggested the need for further investigation of different exercise protocols and intervention intensities with larger sample sizes and more specific outcome measures (17). Since the publication of these systematic reviews, additional clinical trials of exercise on CIPN have been published.

Therefore, we aimed to update the systematic reviews of randomized controlled trials (RCTs) included in the ASCO clinical guidelines by incorporating additional recently published studies evaluating the effect of exercise on the mitigation of CIPN symptoms with an additional focus on the type of exercise program. Since CIPN symptoms and their incidence rate vary by assessment measurements (20), a consensus has been reached that clinical trials investigating CIPN should include an assessment of QOL as an indicator that best reflects the patient’s life state considering various complex symptoms (21). We conducted a meta-analysis with QOL as a primary outcome, and specific outcomes of CIPN symptoms were also analyzed to capture changes in CIPN symptoms associated with exercise interventions. To focus on the details and diversity of exercise intervention programs, we conducted a network meta-analysis (NMA) to determine which exercise interventions were most effective in improving QOL.

## 3 Method

### 3.1 Study design and protocol

We conducted a systematic review and meta-analysis, which followed the guidelines of the Preferred Reporting Items for Systematic Reviews and Meta-Analyses (PRISMA) statement (22). The review protocol was prospectively registered with the University Hospital Medical Information Network Clinical Trials Registry (UMIN000045558). Ethical approval and informed consent were waived considering the nature of the study. This study was conducted as a part of the Japanese clinical practice guidelines on CIPN and was based on a clinical guideline previously published by ASCO (10). Therefore, in the scope of guideline development, the choice was made to include all randomized controlled trials comprising the ASCO systematic review as part of the current analysis (23, 24).

### 3.2 Literature search and data sources

In accordance with the guideline committee policy, a systematic literature search targeted studies published after September 2019 because a previous systematic literature search had been conducted in earlier clinical guidelines published by ASCO (10). A systematic literature search was performed in PubMed on April 20, 2023, to identify RCTs published between September 1, 2019, and April 20, 2023, that evaluated the effects of exercise on CIPN and included more than 5 patients in the exercise group. Interventions specialized only for the upper or lower extremities were excluded. Table S3 presents the search terms used in the publication search strategy. The literature search in PubMed was restricted to articles published in peer-reviewed journals and written in English. In addition to the systematic literature search, we evaluated studies manually obtained primarily by reviewing previously reported clinical guidelines and systematic reviews (10, 11, 13, 17, 25). Two authors independently screened each study for eligibility, and disagreements among reviewers were resolved through discussion.

### 3.3 Data extraction

Two reviewers (SY and NN) independently collected the following information from each included study using a predefined data extraction form: (1) general information about the study (author, year of publication, country, and study design); (2) sample size; (3) patient characteristics (age, sex, cancer type, anticancer drugs that could induce peripheral neuropathy, and the presence of baseline CIPN); (4) exercise type, intensity, duration and frequency; (5) examined outcomes, including CIPN; (6) timing of assessment of outcomes; (7) adverse effects related to the intervention; and (8) enrollment rate, completion rate of the study, and adherence to the intervention. The outcome measures were grouped into six categories referring to a MASCC book (25) as follows: (1) QOL (primary outcome); (2) patient-reported CIPN; (3) pain; (4) clinical assessments of CIPN signs; (5) balance measures; and (6) physical functional assessments. The enrollment rate was defined as the proportion of screened persons who were randomly assigned. The completion rate was defined as the proportion of randomly assigned persons for whom a final analysis was performed. The definition of adherence to exercise intervention followed the individual studies. Discrepancies between the reviewers were resolved through discussion or by a third reviewer (HA).

### 3.4 Quality assessment

Two reviewers (SY and NN) independently assessed the risk-of-bias using version 2 of the Cochrane risk-of-bias tool for randomized trials (RoB 2) (26). All disagreements among reviewers were resolved through discussion. Risk-of-bias assessments were summarized in a traffic-light plot using Risk-of-bias VISualization (robvis) (27). We could not assess the risk of bias, such as heterogeneity among the studies or publication bias, across the studies because many studies included outcomes of different types of measurements and did not specify a primary outcome variable. A funnel plot was used to check for the presence of publication bias.

### 3.5 Data synthesis and meta-analyses

We conducted a meta-analysis on outcome data regarding QOL (primary outcome), patient-reported CIPN symptoms, pain, and collected postintervention or a time point similar to postintervention. We included the studies that reported the data able to calculate the mean and standard deviation at each point. The standardized mean difference (SMD) was calculated due to different scales used across the study outcome measures. Confidence intervals were calculated as a measure of precision for the SMD estimates. If data were unavailable, we contacted the authors and used all data provided by the authors after contact. If a trial was conducted with multiple parallel arms including different types of exercise, we compared the outcomes by combining the arms against a control. The overall mean effect sizes were estimated using random-effects models. Heterogeneity was evaluated with chi-squared tests and *I*-square tests. We used Review Manager (RevMan, Version 5.4., The Cochrane Collaboration, 2020) for the meta-analysis.

### 3.6 Network meta-analysis of exercise intervention effectiveness and QOL

A NMA was conducted to assess the comparative effectiveness of the different exercise interventions in each trial, and the results represent estimates of the relative effects between any given intervention pair in the NMA of QOL. The NMA was conducted using the "BUGSnet" package in R and was based on Bayesian methods (28). We specified 100,000 iterations with 10,000 burn-ins and 10,000 adaptations based on random- and fixed-effects models and compared their accuracies; we employed the model with the lowest deviance information criterion (DIC) and used Bayesian Markov chain Monte Carlo modeling. The treatments were ranked according to cumulative rank curve under surface (SUCRA) probabilities.

## 4 Results

### 4.1 Overview of the included literature and a flow diagram

A total of 1640 studies that were identified for screening after applying the search terms described in Table S3 were screened, and 4 studies (24, 29-31) were included in the final analysis. In addition, the 2 studies (23, 32) included in the update to the ASCO guideline (10) and 6 additional studies (33–38) referred to in previous systematic reviews (11, 13, 17, 25) were included. In total, 12 randomized controlled trials were included in the qualitative synthesis (Figure 1).

**Figure 1.**
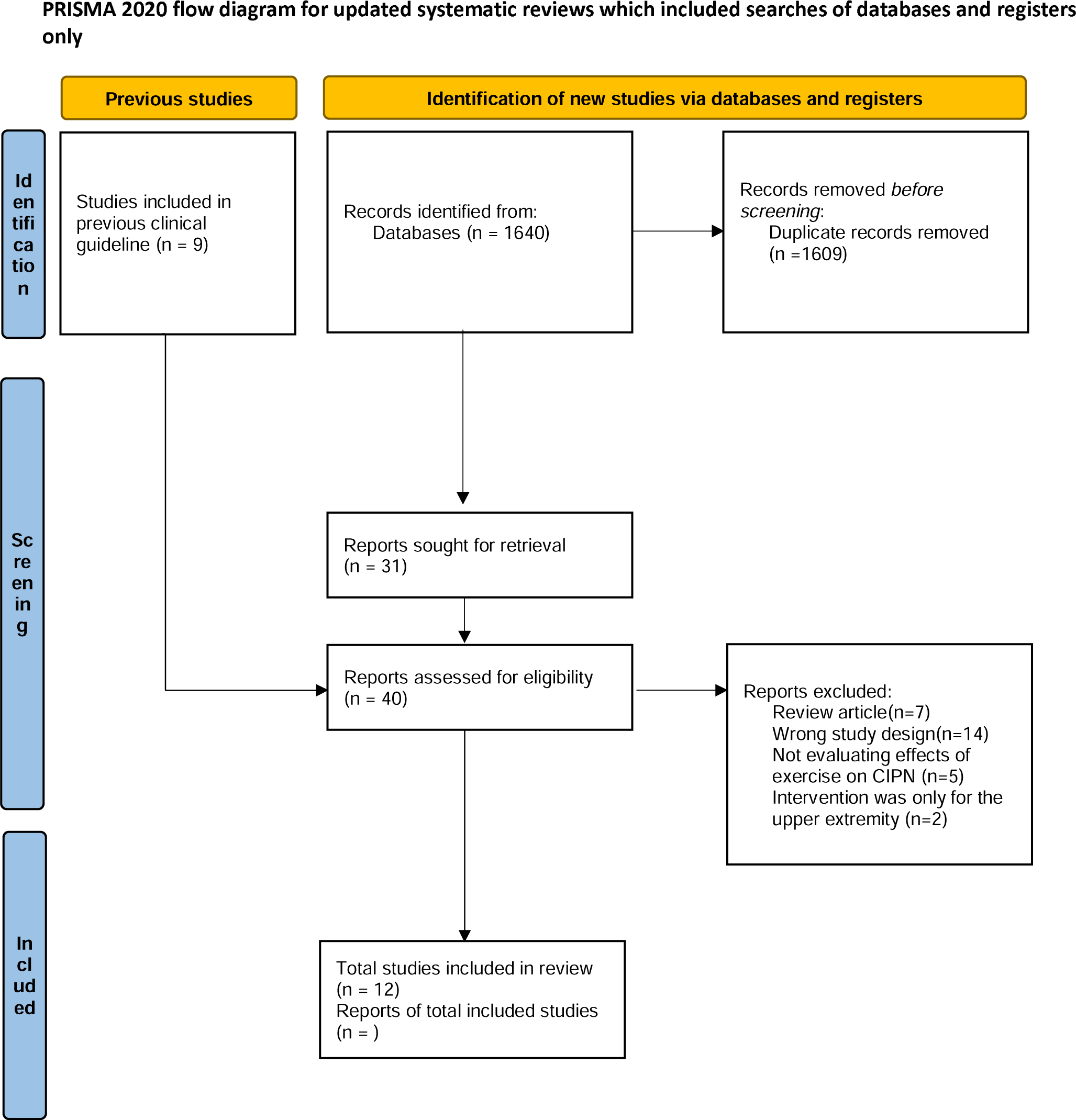
PRISMA flow diagram.

Details on these studies, such as patient disease diagnosis, exercise intervention, and outcome measures, are provided in Table S1.Two-arm (exercise vs. control) study designs were used in 8 studies (23, 24, 31-36); 3-arm designs were used in 2 studies (29, 30); and 4-arm designs were used in 2 studies (37, 38). The control conditions included usual care, education only, and different types of nonpharmacological interventions (Reiki, meditation, cold application). All the included studies aimed to assess the efficacy of exercise on CIPN. Intention-to-treat analysis was conducted in 5 studies (23, 24, 30, 31, 38), and per-protocol analysis was conducted in 2 studies (29, 33). In the remaining 5 studies (32, 34-37), it was unclear which analysis was conducted or how missing data were treated.

### 4.2 Risk-of-bias assessment

The risk of bias was assessed by funnel plots (Figure S1), which were asymmetric and biased toward positive outcomes, and by the RoB 2 tool (Figure S2). For randomization, 5 studies (24, 30, 32, 35, 36) were identified as having a high risk or some concerns for risk-of-bias, as they did not report detailed information about randomization and/or concealment, and one study clearly described that randomization was not masked (37). For deviation, all 12 studies were evaluated to have a high risk of bias. For missing outcome data, 11 studies were identified as high risk or with some concerns for risk-of-bias due to participants dropping out during the study (23, 24, 29–37). For measurement of the outcomes, 10 out of 12 studies were evaluated to have a high risk-of-bias because outcome assessors were often involved in the studies and were aware of the intervention (23, 24, 29, 30, 32-34, 36-38). Finally, for selection in the reported results, 3 studies were evaluated to have a high risk-of-bias because of discrepancies in the description between the methodology and reported results (24, 35, 38).

### 4.3 Study and patient characteristics

The sample sizes of the included studies ranged from 22 to 456 patients. In 6 studies (23, 24, 30, 34, 36, 37), the presence of CIPN before randomization was the eligibility criterion for the respective studies, while in 1 study (29), the absence of CIPN was the eligibility criterion. Patients with breast and gastrointestinal cancers formed the largest population. In all included studies, neurotoxic drugs were cytotoxic agents, and patients with CIPN caused by immune checkpoint inhibitors were excluded.

### 4.4 Intervention details

The types of exercise interventions consisted of balance training (9 studies) (23, 24, 29, 30, 33-36, 38), strength training (7 studies) (23, 24, 29, 30, 32, 33, 35), aerobic training (4 studies) (23, 31–33), and yoga (37). Balance training interventions mainly consisted of standing balanced on an unstable platform under prescribed rules. Seven studies used a combination of different types of exercises (23, 24, 29, 30, 32, 33, 35). In 4 studies (24, 30, 32, 36), the intervention was conducted at home, largely unsupervised, with participants self-reporting their adherence. In 5 other studies (23, 33, 34, 37, 38), the intervention was conducted in the clinic, hospital or sports center, mostly under basic supervision, with an objective assessment of adherence. In one study, participants could choose the place where they would exercise (29). The length of the interventions ranged from 4 to 36 weeks in 11 studies (23, 24, 30–38). In 1 study (29), the intervention continued during chemotherapy, and the intervention duration was not decided beforehand. As shown in Table S(1, the frequency of exercise differed between the studies, ranging from 2 times a week to daily. Likewise, the length of time in which participants were engaged in exercise varied in each study. Most exercise interventions required less than an hour for each session. The intensity of aerobic exercise was adjusted based on the participants’ heart rate in all 3 studies in which the intervention included aerobic exercise (23, 32, 33). Moreover, the intensity was adjusted based on individual capacity (23, 29, 30, 32, 33, 35) in 6 of the 7 studies in which the intervention included strength training. For other types of intervention, the intensity was maintained at a fixed level.

### 4.5 Outcome measurement tools

The primary outcome measures of each study are described in Table S1. Patient-reported CIPN was the most frequent outcome measured (9 studies) (23, 24, 29–32, 34, 37, 38), and QOL was assessed in 8 studies (23, 24, 29, 31, 33, 35, 37, 38). Only one study evaluated the chemotherapy completion rate (29), but none of the studies assessed outcomes related to chemotherapy, including chemotherapy dose received or survival (Table S2).

### 4.6 Effectiveness of exercise on patient-reported outcomes of QOL

Two studies (24, 33) reported a significant intervention effect on QOL. Two studies assessed QOL using the European Organization for Research and Treatment of Cancer core quality of life questionnaire (EORTC-QLQ-C30) (39), and one study assessed QOL using FACT-Taxane (40). The other studies (23, 29, 35, 37, 38) did not detect significant differences in QOL and were assessed with the EORTC-QLQ-C30 or the Functional Assessment of Cancer Therapy-General (FACT-G) questionnaire (41). The synthesized data from 5 studies (23, 24, 29, 31, 37) revealed that exercise significantly improved QOL (SMD, 0.45; 95% CI = 0.12 to 0.(78, P = 0.(008, *I*^2^ = 31%) (Figure 2A).

**Figure 2.**
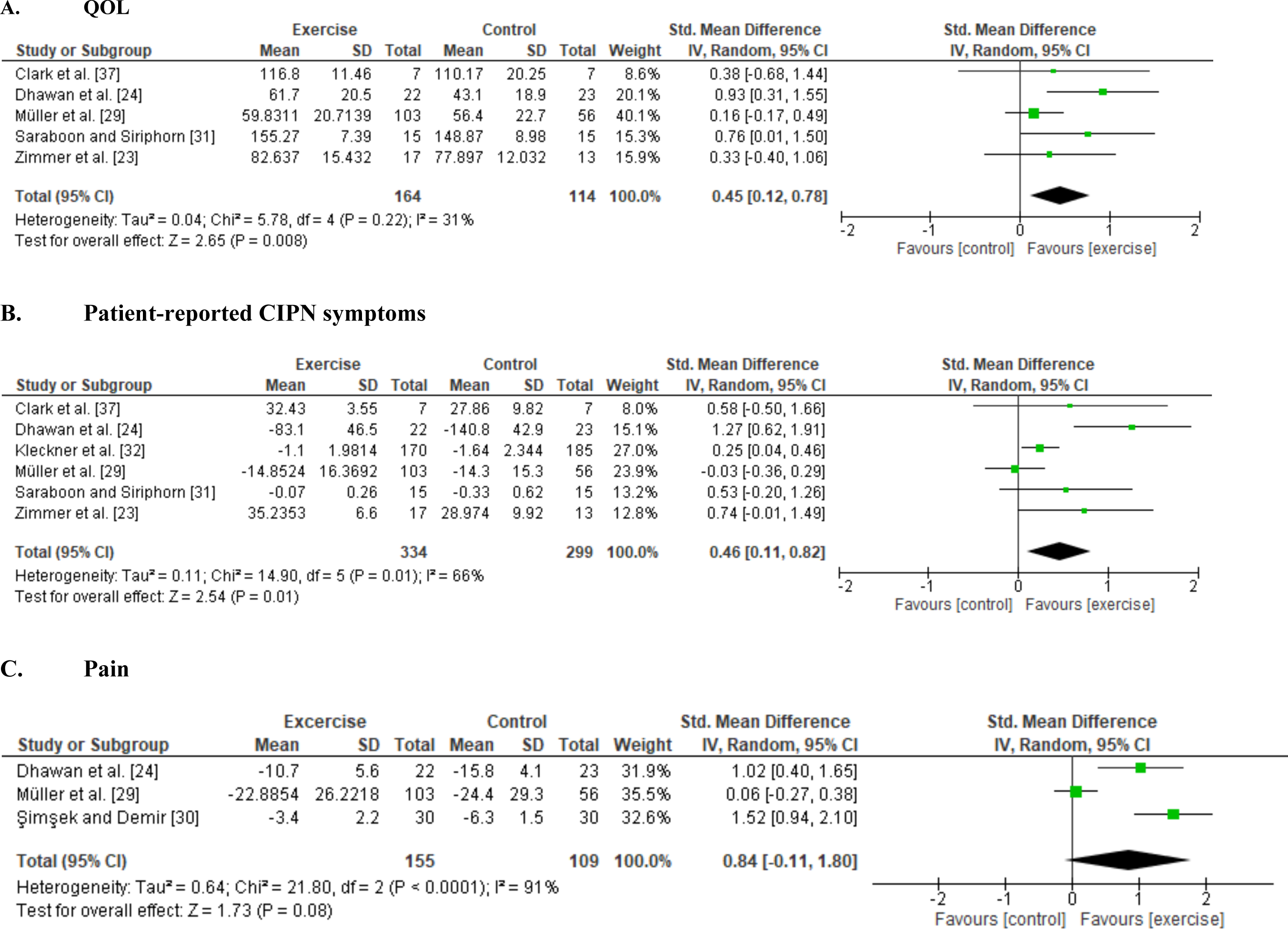
Forest plots of the effects of exercise on patient-reported outcomes. (A) Quality of life (QOL), (B) Patient-reported CIPN symptoms, (C) Pain.

### 4.7 Effectiveness of exercise for patient-reported CIPN symptoms

Four studies (23, 24, 30, 32) reported significant intergroup differences in patient-reported symptoms related to CIPN, whereas four studies (29, 31, 34, 38) did not detect significant differences. One study compared pre- and postintervention data and reported that neuropathic symptoms significantly worsened only in the control group (37). Although three studies (23, 37, 38) assessed patient-reported CIPN symptoms using the Functional Assessment of Cancer Therapy/Gynecologic Oncology Group Neurotoxicity (FACT-GOG-NTX) questionnaire (42), the results were inconsistent. The six studies (23, 24, 29, 31, 32, 37) included in the meta-analysis showed a significant intervention effect on patient-reported CIPN symptoms (SMD, 0.46; 95% CI = 0.11 to 0.(82, P = 0.(01, *I*^2^ = 66%) (Figure 2B). Fear of falling was not included as a patient-reported CIPN symptom in the meta-analysis because of high heterogeneity (29, 34).

### 4.8 Adverse events related to exercise

Nine studies (23, 24, 29, 32-(36, 38) reported adverse events; however, of these, only Müller et al. (29) reported adverse events related to the exercise programs, in which pain, fatigue and dizziness occurred in 21–25% of patients in the exercise group (29).

Both studies where pain was measured using pain scales (24, 38) reported a significant reduction in pain in the exercise group compared to the control group. There were no significant differences in pain scores on the Chemotherapy-Induced Peripheral Neuropathy Assessment Tool (CIPNAT) (30) or the EORTC-QLQ-C30 (29, 33, 38). A forest plot showed no significant difference between the exercise groups and control groups, but a trend towards a favorable effect of exercise was observed (SMD, 0.84; 95% CI = −0.11 to 1.(80, P = 0.(08, I^2^ = 91%) (Figure 2C).

### 4.9 Clinical assessments of CIPN post-exercise intervention

Clinical assessments of CIPN symptoms were significantly improved by exercise in 2 out of 3 studies (31, 33, 38). The beneficial effects of the intervention included improvements in peripheral deep sensitivity (33, 38) and Achilles tendon and patellar tendon reflexes (38). Significant intervention effects on balance ability were observed in all relevant studies (23, 29, 33–36). However, outcomes related to balance involved multiple measures and a wide range of signs. Mixed results were observed for physical function. Four studies reported benefits to muscle strength (23, 29, 35) and SPPB (31), while three studies did not show beneficial effects (33, 34, 36).

### 4.10 Enrollment, completion rate and adherence

Patient enrollment rates were described in 7 studies (24, 29-(31, 34, 35, 37) and ranged from 25 to 100% (Table S4). Two studies (24, 35) described the reasons for the decrease in the enrollment rate, including lack of interest, personal reasons, and unfulfilled eligibility criteria. Completion rates were described in all studies and ranged from 64 to 100%. Six studies (23, 32, 34, 36–38) described the reasons for the decrease in the completion rate, including death, disease progression, and physical, psychological, and personal reasons. Adherence to the interventions was described in 8 studies (23, 24, 29, 31–33, 36, 38) and ranged from 49 to 98%. Two studies (29, 36) described the details of the reasons for the decrease in adherence, including side effects of anticancer treatment, time constraints and motivation.

### 4.11 Network meta-analysis of exercise interventions on QOL

A NMA was conducted to compare the effectiveness of each exercise intervention on QOL and included each trial arm for which a QOL index was available (23, 24, 29, 37) (Figure 3A. network graph). The fixed-effects model was selected for this data set.

**Figure 3.**
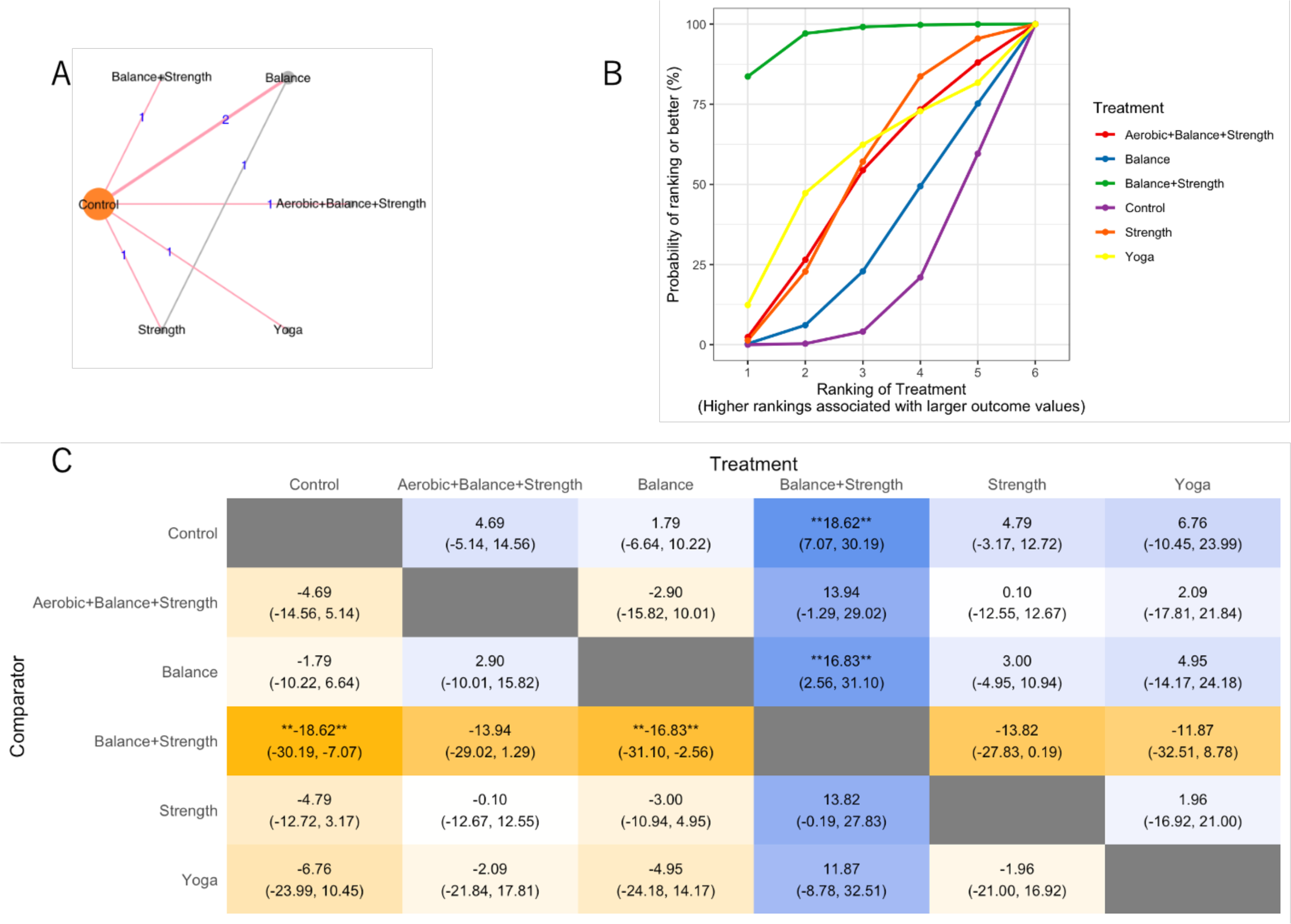
Network meta-analysis(A) network graph, (B) SUCRA plot, (C) League Table Heatmap. The values in each cell show the relative treatment effect and 95% credible intervals of the treatment on the top, compared to the treatment on the left. The double asterisks: statistical significance.

In the SUCRA plot (Figure 3B), the curve associated with the combination treatment balance and strength training had the highest probability of being ranked first among treatments, suggesting that it is most likely to result in improved QOL scores. The model also generates effect estimates. The difference between balance and strength training and the other treatments was larger than that of the control and balance training (Figure 3C).

## 5 Discussion

The meta-analyses indicated that exercise was effective in improving QOL scores and patient reported CIPN symptoms. The results of the NMA on specific exercise interventions showed that the combination of balance and strength training provided the greatest benefit to QOL over the control. Severe adverse events regarding exercise were reported to be rare, and the meta-analysis showed that pain status after the exercise intervention tended to be positive in the intervention group. Considering these findings, exercise interventions that include strength training confer the greatest benefit for CIPN.

Our analysis with updated trials indicated that exercise positively affected QOL, consistent with previous systematic reviews that showed that exercise resulted in an improvement in QOL scores (SMD: 14.(62, 95% CI, 6.03–3.20) (19, 43). A similar meta-analysis of studies on the severity of CIPN and peripheral deep sensitivity showed positive results, as in our research on QOL(44). These exercise interventions differed among the 12 studies included in this systematic review. Concerning intervention programs, a daily home-based program combining balance and strength training was suggested to have the greatest improvement in QOL compared with that of the control group, but the differences were not statistically significant in the NMA. While most of the other studies included in this analysis used an exercise frequency of 2–3 times per week, the trial that used the Strength + Balance exercise intervention had a daily home-based intervention, suggesting that a high frequency of specialized exercise was implemented. These results are similar to those of a clinical trial that compared different doses and modes of exercise for breast cancer patients who underwent chemotherapy and showed that compared with standard volumes, a higher volume of aerobic or combined exercise may better manage declines in physical functioning and worsening symptoms (45). Although exercise relieves general chronic pain, it causes acute muscle pain (46); however, the results of this study suggest that strength exercise does not cause pain in patients with CIPN. In cancer patients, intrapersonal factors (functional status or pain, interest in exercise, and beliefs about the importance of exercise) are related to low physical activity (47). However, it is recommended for general cancer survivors to avoid physical inactivity and perform at least 150–300 minutes of moderate-intensity exercise or 75 minutes of high-intensity exercise, or an equivalent combination, per week, with at least two days of resistance training (muscle strengthening exercise) (15, 48). Implementing these common exercise recommendations for cancer survivors is expected to improve pain and remove the barriers to exercise. The results of this NMA suggest that a program that adds balance training to these exercise recommendations may help improve the QOL of CIPN patients.

There are two limitations of this research. First, the included studies are predominantly small exploratory studies with limited information, and it is difficult to exclude the possibility that study biases may have led to positive results. It is recommended that the effect-size metrics selected for the meta-analysis be comparable and reliable, but none of them are perfect in this report. The funnel plot results were asymmetric and biased towards positive outcomes, which indicates selection bias. There are several reasons why bias may have been observed, including protocol deviations due to individual differences in exercise performance, self-reported outcome measures, nonblinded interventions, and the possibility of survival bias among patients with high adherence to the described study protocols. In addition, CIPN symptoms are influenced by the type and schedule of chemotherapy, but no study has established or described those factors in detail. Therefore, differences in the target population may have influenced the results. The consideration of these sources of potential bias is important when interpreting the results of these exercise interventions. Furthermore, isolating an actual intervention effect from a placebo effect is difficult to achieve, as participation in a study itself may lead to improvements in motivation and physical and mental function. Altogether, this highlights the clinical importance of conducting a meta-analysis and NMA to consider the totality of evidence that supports the claim for exercise in the management of CIPN.

Second, many trials did not specify whether the tested intervention was intended to prevent or treat CIPN. Since symptoms of CIPN can occur in either limb, interventions should ensure that assessments are made at the site of CIPN onset. However, it is difficult to predict the CIPN site and complete a functional assessment and intervention by the site in the research framework, especially in prevention research. For future research, it is important to design exercise interventions with multiple effects and evaluation methods, such as large-scale clinical trials with subgroup analysis, in advance.

Based on the results of this study, it is expected that not only would a reduction in CIPN symptoms improve QOL but also that an improvement in basic physical functions, such as underlying physical fitness, has a synergistic effect of enabling a more active lifestyle and improving QOL. The present NMA could not show the significant effects but suggested that a combination of balance and strength training, which require the supervision of exercise specialists and the creation of an appropriate environment. The implementation of active lifestyles is recommended (15), and they can be easily introduced into the daily lives of cancer patients. However, our NMA results suggest that high-frequency combined strength and balance exercise interventions are preferable for reducing CIPN, which requires more specialized guidance than general lifestyle physical activities.

## 6 CONCLUSION

Our results indicate that the exercise interventions implemented in the RCTs are effective in improving QOL and CIPN symptoms. While increasing physical activity would enhance general QOL and aerobic exercise is observed to be an effective approach, the combination of balance and strength training provides the great benefits in CIPN patients. Our meta-analysis results strengthen the evidence that exercise benefits the QOL of CIPN patients. Furthermore, daily exercise with a combination of balance and strength training would promise for improving QOL.

Further research on how to implement a system incorporating more specialized and intense exercise in this patient population is needed to achieve exercise implementation based on an assessment of balancing ability and efficient strength training on a continuous basis.

## 8 Author Contributions

All authors contributed to the idea for the article and the search strategy planning. NN, SY, AO and HA conducted screening of the articles. NN and SY performed data extraction and quality assessment. AO and HA contributed to review of the data. NN, SY, and AH drafted the manuscript. AO and HA critically revised the drafted manuscript. All authors have reviewed and approved the submitted version of the manuscript.

## 10 Acknowledgments

We would like to thank the authors of the original papers who provided us with information for this study.

## Supplementary materials

**Table S3.** The detail of search strategy in this study

**Table S4.** Enrollment, completion rate and adherence

## 1 Data Availability Statement

The data that support the findings of this study are available from the corresponding author upon reasonable request.

