## Supplementary material for "Exercise intervention for the management of chemotherapy-induced peripheral neuropathy: A systematic review and network meta-analysis": TableS1 and Table S2

**Table S1.** Overview of included studies

| Author and Year | Exercise Type | Patients | Details of Exercise | Main Outcome Measures † |
| --- | --- | --- | --- | --- |
| Streckmann et al. | Aerobic | Lymphoma, mixed chemo types | ~1 hr/d, 2 d/wk, 36 wk | Baseline, after 12, 24, and 36 wk |
| 2014 [33] | Balance | Unclear baseline CIPN presence | In the clinic, supervised | **EORTC QLQ-C30** |
|  | Strength | N = 61 (total)  N = 30 (exercise) | Aerobic: warm-up on a bicycle dynamometer (60–70% HR max), 10–30 min on a treadmill or bicycle dynamometer (70–80% HR max) at the end of the session | Vibration |
|  |  | 2 arms | Balance: four postural stabilization exercises, progressively increasing exercise difficulty as well as surface instability (20 s exercise +20 s rest for 3 sets, 1 min rest between different exercises) | Balance control (on static/dynamic surface) |
|  |  |  | Strength: four different resistance exercises performed with Thera-Bands, at maximum resistance. | Incremental step test |
| Schwenk et al. | Balance | Mixed cancers, chemo types | 45 min/d, 2 d/wk, 4 wk | Baseline and post- intervention |
| 2016 [34] |  | Established CIPN | In the clinic, unsupervised | FES-1 (fear of falling) |
|  |  | N = 22 (total)  N = 11 (exercise) |  | **Balance** (feet close/semi tandem, eyes open/feet close, eyes closed) |
|  |  | 2 arms |  | Gait-speed variability |
| Vollmers et al. | Balance | Breast cancer, during paclitaxel treatment | 2d/wk, during chemotherapy and for 6 wk after chemotherapy | Baseline, at the last dose of chemotherapy and at the 6-wk follow-up |
| 2018 [35] | Strength | Unclear baseline CIPN presence | Balance: no dose specified | EORTC QLQ CIPN 20 EORTC QLQ-C30 EORTC-BR23 |
|  |  | N = 43 (total)  N = 21 (exercise) | Strength: 6 exercises, 2 sets of 20 reps, RPE: 13-15 (moderate intensity) | **Posturometry sway area Fullerton Advanced Balance Scale scores** |
|  |  | 2 arms |  | Hand dynamometer Chair rising test |
| Zimmer et al. | Aerobic | Gastrointestinal cancers, oxaliplatin treatment | 60 min/d, 2d/w, 8wk | Baseline, postintervention(8 wk), and12 wk |
| 2018 [23] | Balance | Established CIPN | In a sports center, supervised | **FACT/GOG-TOI** FACT/GOG-NTX |
|  | Strength | N = 30 (total)  N = 17 (exercise) | Aerobic: cross-trainer, ergometer or walking; 10 min, 60-70% HRmax | GGT-Reha |
|  |  | 2 arms | Balance: 10 min | Strength: e1RM 6MWT |
|  |  |  | Strength: 5 exercises; 20 min; 60-80% of e1RM; Borg CR10 scale level: 6 |  |
| Kleckner et al. | Aerobic | Mixed cancers, chemo types | Daily, 6wk | Baseline and post- intervention |
| 2018 [32] | Strength | Most patients reported mild baseline CIPN | Home based, unsupervised | **Numbness (NRS) Tingling (NRS) Hot/coldness in hands/feet (NRS)** |
|  |  | N = 456 (total)  N = 231 (exercise) | Aerobic: walking; 60-85% of heart rate reserve |  |
|  |  | 2 arms | Strength: resistance exercise; 3-5 RPE |  |
| Stuecher et al. | Aerobic | Gastrointestinal cancers, mixed chemo types | 12 wk | Before chemotherapy, after 2 cycles, after 12 weeks |
| 2019 [36] |  | Unclear baseline CIPN presence | Home based, unsupervised | Vibration |
|  |  | N = 44 (total)  N = 22 (exercise) | 150 min, moderate walking per week | Postural sway |
|  |  | 2 arms | Borg’s self-rating of RPE of 11-13 | **SPPB** Gait speed Lower-extremity muscle strength |
| Clark et al. | Yoga* | Mixed cancers, platinum treatment | 60 min/w, 6wk | Baseline and post-intervention |
| 2012 [37] |  | Established CIPN | In the clinic, supervised | FACT-GOG-Ntx Brief Symptom Inventory – 18 |
|  |  | N = 36 (total)  N = 9 (exercise) | Low intensity |  |
|  |  | 4 arms |  |  |
| Streckmann et al. | Balance | Mixed cancers, chemo types | 2d/wk, 6wk | Baseline and post-intervention |
| 2019 [38] |  | Established CIPN | In the clinic, supervised | FACT GOG-Ntx EORTC-QLQ-C30 Pain-DETECT |
|  |  | N = 40 (total)  N = 20 (exercise) | Balance: on progressively unstable surfaces; 4 exercises; 3 sets of 20 s | **Achilles tendon reflex Deep sensitivity Patellar tendon reflex Light-touch perception Sense of position** Nerve conduction velocity and amplitude |
|  |  | 4 arms | Whole-body vibration: stand on a vibration platform; 4 sets of 30-s to 1-min | Balance control |
|  |  | Balance/Whole-body vibration/Control/Healthy control |  | **Lower-leg strength** Gait speed Postural sway |
| Dhawan et al. | Balance | Mixed cancers, carboplatin + paclitaxel | 30 min/d, daily, 10 wk | Baseline and post-intervention |
| 2020 [24] | Strength | Established CIPN | Home based, unsupervised | CIPNAT **LANSS EORTC-QLQ** |
|  |  | N = 45 (total)  N = 22 (exercise) |  |  |
|  |  | 2 arms |  |  |
| Müller et al. | Balance | Mixed cancers, chemo types | During chemotherapy | Baseline, post- intervention and 3 and 6 wk after the intervention |
| 2021 [29] | Strength | No baseline CIPN | Balance: 35 min/d, 3/wk, at home (unsupervised) or in the hospital (supervised) | **TNS** EORTC QLQ-CIPN 15 EORTC QLQ-C30 FES-1 (fear of falling) |
|  |  | N = 170 (total)  N = 112 (exercise) | Strength: 45 min/d, 2/wk, machine-based & 15 min/d, 1/wk, home-based | Nerve conduction studies |
|  |  | 3 arms |  | Postural control (Balance) |
|  |  | Balance/Strength/Control |  | Lower-extremity score |
|  |  |  |  | Chemotherapy completion rate |
| Şimşek and Demir | Balance | Breast cancer, mixed chemo types | 15-30 min/d, 5d/wk, 12 wk | Baseline and post-intervention |
| 2021 [30] | Strength | Established CIPN | Home based, supervised | **CIPN Assessment Tool** |
|  |  | N = 90 (total)  N = 30 (exercise) | Balance: 4 exercises |  |
|  |  | 3 arms | Strength: 7 exercises |  |
|  |  | Exercise/cold application/control | 10 reps/set for the first 3 wk, then 20 reps/set for 3 wk, and then 30 reps/set for 3 wk |  |
| Saraboon and Siriphorn | Balance | Ovary or cervix cancer, paclitaxel regimens | During chemotherapy | Baseline, during intervention (4 wk) and postintervention (6 wk) |
| 2021 [31] |  | No baseline CIPN | 60 min/d, 2d/wk, 6 wk | FAB |
|  |  | N = 30 (total)  N = 15 (exercise) |  | MDNS |
|  |  | 2 arms |  | FACT-Taxane |
|  |  |  |  | SPPB |

†, Primary endpoint is written in bold and underlined. *, Relaxation-based hatha yoga consisting of weekly lessons and daily self-practice.

CIPN: chemotherapy-induced peripheral neuropathy, CIPNAT: Chemotherapy-Induced Peripheral Neuropathy Assessment Tool, DASH: Disability of the Arm, Shoulder, and Hand, e1RM: estimated one-repetition maximum, EORTC QLQ: the European Organization for Research and Treatment of Cancer core quality of life questionnaire, FAB: Fullerton Advanced Balance, FACT-GOG: Functional Assessment of Cancer Therapy/Gynecologic Oncology Group, TOI: Trial Outcome Index, Ntx: Neurotoxicity, FACT-Taxane: Functional Assessment of Cancer Therapy-Taxane, FES-1: falls efficacy scale-1, HR: heart rate, LANSS: Leeds Assessment of Neuropathic Symptoms and Sign, MDNS: Michigan Diabetic Neuropathy Score, NPRS: Numeric Pain Rating Scale, NRS: number rated scale, RPE, rate of perceived exertion, S-LANSS: Self-report version of Leeds Assessment for Neuropathic Symptoms and Signs, SPPB: short physical performance battery, TNS: total neuropathy score

**Table S2.** Summary of main results regarding the effect of exercise on outcomes

| Study | Quality of Life (QOL) | Patient-reported CIPN | Pain | Clinical assessments of CIPN signs | Balance measures | Physical functional assessments |
| --- | --- | --- | --- | --- | --- | --- |
| Intergroup comparison | | | | | | |
| Streckmann et al. 2014 [33] | **Improvement in QOL within the first 12 weeks**  Δ_T1–T0_ (Mean): IG 9.1, CG -6.15, P = 0.028 |  | No significant difference | **Reduced peripheral deep sensitivity after 36 weeks**  IG 87.5% (symptom diminished), CG 0%, P < 0.001 | **Reduced sway paths after 36 weeks**  Δ_T3–T0_: static left P = 0.035, dynamic left P = 0.007, dynamic right P = 0.045  **Improvement in failed attempts**  Δ_T3–T0_: static left P = 0.024, dynamic left P = 0.014, dynamic right P < 0.001  **Improvement in time to regain balance after 36 weeks**  Δ_T3–T0_ (Median): IG -0.26, CG 0.2, P = 0.045 | No significant difference |
| Schwenk et al. 2016 [34] |  | No significant difference |  |  | **Reduced sway of hip, ankle, and center of mass (CoM) post intervention**  Feet close – eyes open P = 0.010–0.022 (except anterior-posterior CoM sway)  Semitandem – eyes open  P = 0.008–0.035 (except ankle sway) | No significant difference |
| Vollmers et al. 2018 [35] | No significant difference |  |  |  | **Smaller sway area**  Monopedal stance  T1 (after the last dose) (left and right) both P < 0.001, T2 (6 weeks follow-up) (left) P = 0.003, T2 (right) P<0.01  Bipedal stance  P = 0.039  **Improvement in postural stability**  Δ_T1-T0_: IG +1.35, CG -2.84, P < 0.001 | **Less loss of strength in hand dynamometry**  IG 0.60, CG -1.60, P = 0.029 |
| Zimmer et al. 2018 [23] | No significant difference | **Less aggravation of neuropathic symptoms**  Mean (T0/T1[after the intervention]/T2[after 4 weeks follow-up]): IG 33.12/35.24/34,  CG 34.08/28.97/29.43  Δ_T1–T0_: P = 0.002, Δ_T2–T0_: P = 0.015 |  |  | **Improvement in advanced static balance**  Mean (T0/T1/T2):  IG 11.47/13.35/12.47,  CG 11/9.92/9.62  Δ_T1–T0_: P = 0.025, Δ_T2–T0_: P = 0.025 | **Increased muscle strength**  Bench press  Δ_T1–T0_: P = 0.014, Δ_T2–T0_: P = 0.014  Leg press  Δ_T1–T0_: P = 0.001, Δ_T2–T0_: P = 0.011  Lat pulldown  Δ_T1–T0_: P = 0.022, Δ_T2–T0_: P = 0.031 |
| Kleckner et al. 2018 [32] |  | **Less severe CIPN symptoms at postintervention**  Hot/coldness in hands/feet  Coefficient: -0.46, P = 0.045 |  |  |  |  |
| Stuecher et al. 2018 [36] |  |  |  |  | **Improvement in postural sway after 12 weeks**  Δ_T2–T0_ (Mean): IG -58.8, CG 58.7, P = 0.003 (Group effect) | No significant difference |
| Streckmann et al. 2019 [38] | No significant difference | No significant difference | **Reduction of pain**  F(2,25) = 3.575, P = 0.043 | **Improvement in Achilles tendon reflex**  F(2,26) = 4.791, P = 0.017  **Improvement in patellar tendon reflex**  F(2,25) = 4.564, P = 0.020  **Improvement in peripheral deep sensitivity**  F(2,25) = 5.548, P = 0.010 |  |  |
| Dhawan et al. 2020 [24] | **Better QOL after 10 weeks**  Mean: IG 61.7, CG 43.1, P = 0.002 | **Less severe, distressing, or frequent CIPN symptoms after 10 weeks**  Mean: IG 83.1, CG 140.8, P < 0.0001 | **Less neuropathic pain after 10 weeks**  Mean: IG 10.7, CG 15.8, P = 0.001 |  |  |  |
| Müller et al. 2021 [29] | No significant difference | No significant difference | No significant difference | No significant difference | **Keeping average time of standing on one leg with open eyes at 3 weeks after chemotherapy**  Adjusted between-group difference:  Sensorimotor training vs. CG 2.2, P = 0.045  Resistance training vs. CG 2.3, P = 0.023 | **Maintained muscle strength**  Adjusted between-group difference:  Resistance training vs. CG 11.1, P = 0.045 |
| Şimşek and Demir 2021 [30] |  | **Less severe CIPN symptoms after 12 weeks**  Numbness in the hand  Mean: IG 1.9, Cold application 3.0, CG 3.1, P = 0.009  Numbness in the foot  Mean: IG 3.8, Cold application 4.0, CG 5.5, P = 0.009 | No significant difference |  |  |  |
| Saraboon and Siriphorn  2021 [31] | No significant difference | No significant difference |  |  | **Invariant balance performance**  Mean: IG 37.40, CG 34.13, P < 0.01 | **Maintained baseline physical performance**  Mean: IG 11.47, CG 10.67, P = 0.03 |
| Intragroup comparison | | | | | | |
| Clark et al. 2012 [37] | No significant difference | **There were no significant changes in IG whereas neuropathic symptoms worsened in CG.**  Mean, pre vs. post  IG: 30.31 vs. 32.43, P = 0.278  CG: 31.14 vs. 27.86, P = 0.03 |  |  |  |  |

CIPN: chemotherapy-induced peripheral neuropathy, IG: intervention group, CG: control group
