## Supplementary figures and images for "Exercise intervention for the management of chemotherapy-induced peripheral neuropathy: A systematic review and network meta-analysis"

### FigureS1

**Figure S1.**

**A. QOL**

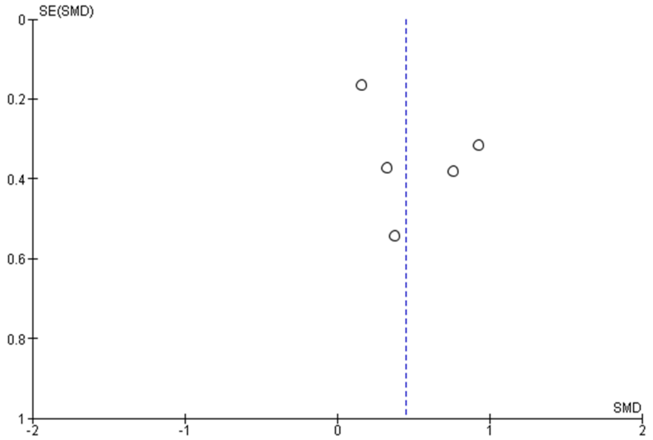


**B. Patient-reported CIPN symptoms**

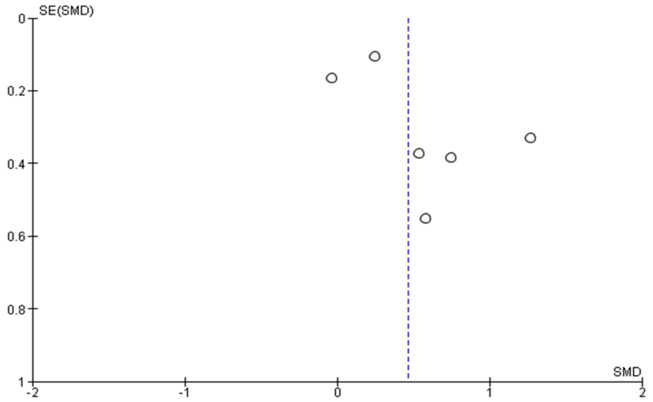


**C. Pain**

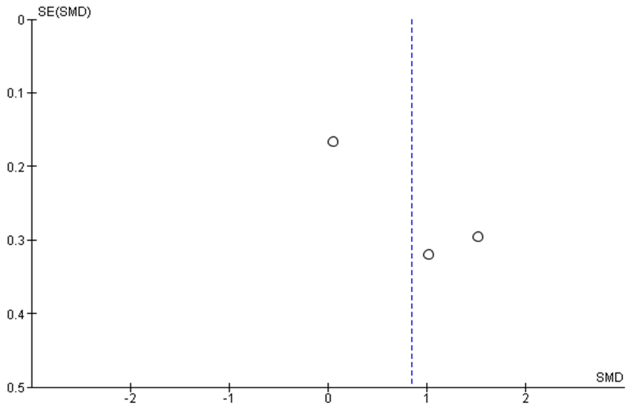
