## Supplementary material for "Exercise intervention for the management of chemotherapy-induced peripheral neuropathy: A systematic review and network meta-analysis": FigureS2

**Figure S2.**

Risk of bias domains

| Study | D1 | D2 | D3 | D4 | D5 |
|---|---|---|---|---|---|
| Streckmann et al. [33] | + | X | X | X | + |
| Schwenk et al. [34] | + | X | X | X | + |
| Vollmers et al. [35] | X | X | X | - | X |
| Zimmer et al. [23] | + | X | X | X | + |
| Kleckner et al. [32] | - | X | X | X | + |
| Stuecher et al. [36] | - | X | X | X | + |
| Clark et al. [37] | X | X | X | X | + |
| Streckmann et al. [38] | + | X | + | X | X |
| Dhawan et al. [24] | - | X | - | X | X |
| Müller et al. [29] | + | X | X | X | - |
| Şimşek and Demir [30] | - | X | X | X | + |
| Saraboon and Siriphorn [31] | + | X | X | + | + |

Domains:
D1: Bias arising from the randomization process.
D2: Bias due to deviations from intended intervention.
D3: Bias due to missing outcome data.
D4: Bias in measurement of the outcome.
D5: Bias in selection of the reported result.

Judgement
X High
- Some concerns
+ Low
